# Genetically predicted plasma levels of amino acids and stroke risk: a Mendelian randomization study

**DOI:** 10.1101/2024.11.07.24316941

**Authors:** Zeheng Li, Yutong Zhang, Hongjie Zhou, Yu Xu, Lulu Sun, Zhen Zhang, Zhongyu Gao, Suyu Wang, Jianqiang Ni, Zhigang Miao

## Abstract

Stroke, including ischemic stroke (IS) and small vessel stroke (SVS), is a major cause of morbidity and mortality globally. The role of amino acids in stroke risk and outcomes is not well understood. This study investigates the causal effects of genetically determined amino acid levels on stroke and its functional outcomes using Mendelian randomization (MR). We analyzed data by single nucleotide polymorphisms (SNPs), Inverse-variance weighted (IVW) and so on. After False discovery rate (FDR) correction, we found that Higher genetically determined levels of CSF glycine (odds ratio [OR] per standard deviation [SD] increase, 1.34; 95% confidence interval [CI], 1.14-1.56; P=2.46×10-4), glutamate (odds ratio [OR] per standard deviation [SD] increase, 1.48; 95% confidence interval [CI], 1.17-1.87; P=9.50×10-4), glutamine (odds ratio [OR] per standard deviation [SD] increase, 1.58; 95% confidence interval [CI], 1.29-1.94; P=1.30×10-5), and phenylalanine (odds ratio [OR] per standard deviation [SD] increase, 1.58; 95% confidence interval [CI], 1.32-1.89; P=7.37×10-7) were associated with increased risks of SVS. Higher phenylalanine (odds ratio [OR] per standard deviation [SD] increase, 1.79; 95% confidence interval [CI], 1.26-2.55; P=1.15×10-3) was linked to increased risks of worse IS functional outcomes (modified Rankin Scale score≥3). These findings suggest amino acids as potential biomarkers and therapeutic targets for stroke.

## Introduction

Stroke remains a significant global health challenge, ranking among the leading causes of death and disability(Katan and Luft 2018). Ischemic stroke (IS), caused by an obstruction in the blood vessels supplying the brain, constitutes the 87% of all strokes(Hui et al. 2024). The identification of modifiable risk factors and underlying mechanisms contributing to stroke incidence and outcomes is crucial for improving prevention and treatment strategies. Emerging evidence suggests that metabolic factors, including amino acid levels, may play a critical role in the pathophysiology of stroke (Phang et al. 2008; Vinknes et al. 2021).

Amino acids, as fundamental building blocks of proteins, are involved in numerous metabolic pathways that influence vascular health and brain function(Polis and Samson 2020; Ling et al. 2023). However, the causal relationships between specific amino acids, especially those in cerebrospinal fluid, and stroke risk and recovery remain inadequately understood. Traditional observational studies are often limited by confounding factors and cannot establish causality (Andrade 2014; Hammerton and Munafò 2021). Therefore, a more rigorous approach is needed to elucidate these potential links.

MR is a robust analytical technique that uses genetic variants as instrumental variables (IVs) to assess the causal effects of exposures on health outcomes(Richmond and Davey Smith 2022; Sanderson et al. 2022). Additionally, a two-sample MR design can significantly expand the scope of MR analysis by utilizing the advantages of two independent sample(Zheng et al. 2017). By utilizing genetic variants associated with amino acid levels, MR can provide insights into whether alterations in amino acid metabolism contribute to stroke risk and functional outcomes following IS, thereby overcoming limitations of conventional epidemiological studies.

This study aims to address critical gaps in our understanding of the role of amino acids in stroke by employing MR analysis. Specifically, it investigates the causal effects of genetically determined circulating amino acid levels and cerebrospinal fluid amino acid levels on the incidence of IS, SVS, and functional outcomes following IS. By identifying amino acids that may influence stroke risk and recovery, this research could reveal novel metabolic targets for therapeutic intervention and risk stratification.

The clinical significance of this investigation lies in its potential to enhance stroke prevention and management. If specific amino acids are found to have causal effects on stroke outcomes, they could serve as biomarkers for early detection, prognosis, and personalized treatment strategies. Moreover, understanding these metabolic pathways may lead to the development of new nutritional or pharmacological interventions aimed at modulating amino acid levels to reduce stroke risk and improve recovery. This study, therefore, seeks to provide a foundational understanding that could inform future clinical practices and public health policies aimed at mitigating the burden of stroke.

## Materials and Methods

### Study design

The present study was reported using the Strengthening the Reporting of Observational Studies in Epidemiology using Mendelian Randomization (STROBE-MR) guideline (Skrivankova et al. 2021). As depicted in Figure 1, we employed a two-sample MR approach to investigate the influence of amino acids on the incidence of stroke and stroke functional outcome. We selected single-nucleotide polymorphisms (SNPs) that reached genome-wide significance (at the genome-wide significance level (P<5.0×10^-6^) and were not in linkage disequilibrium (LD) with other SNPs (r^2^<0.001 within a clumping window of 10000 kb)) for amino acids. Summary genetic data on ischemic stroke and its subtypes were sourced from the Multi-ancestry Genome-Wide Association Study conducted by the International Stroke Genetics Consortium (MEGASTROKE) and GISCOME network, including 6,021 patients of European descent (Malik et al. 2018; Söderholm et al. 2019). All participants in this MR analysis were of European ancestry. The protocol and data collection for the original genome-wide association studies (GWASs) were approved by an ethics committee, and written informed consent was obtained from each participant.

**Figure 1.**
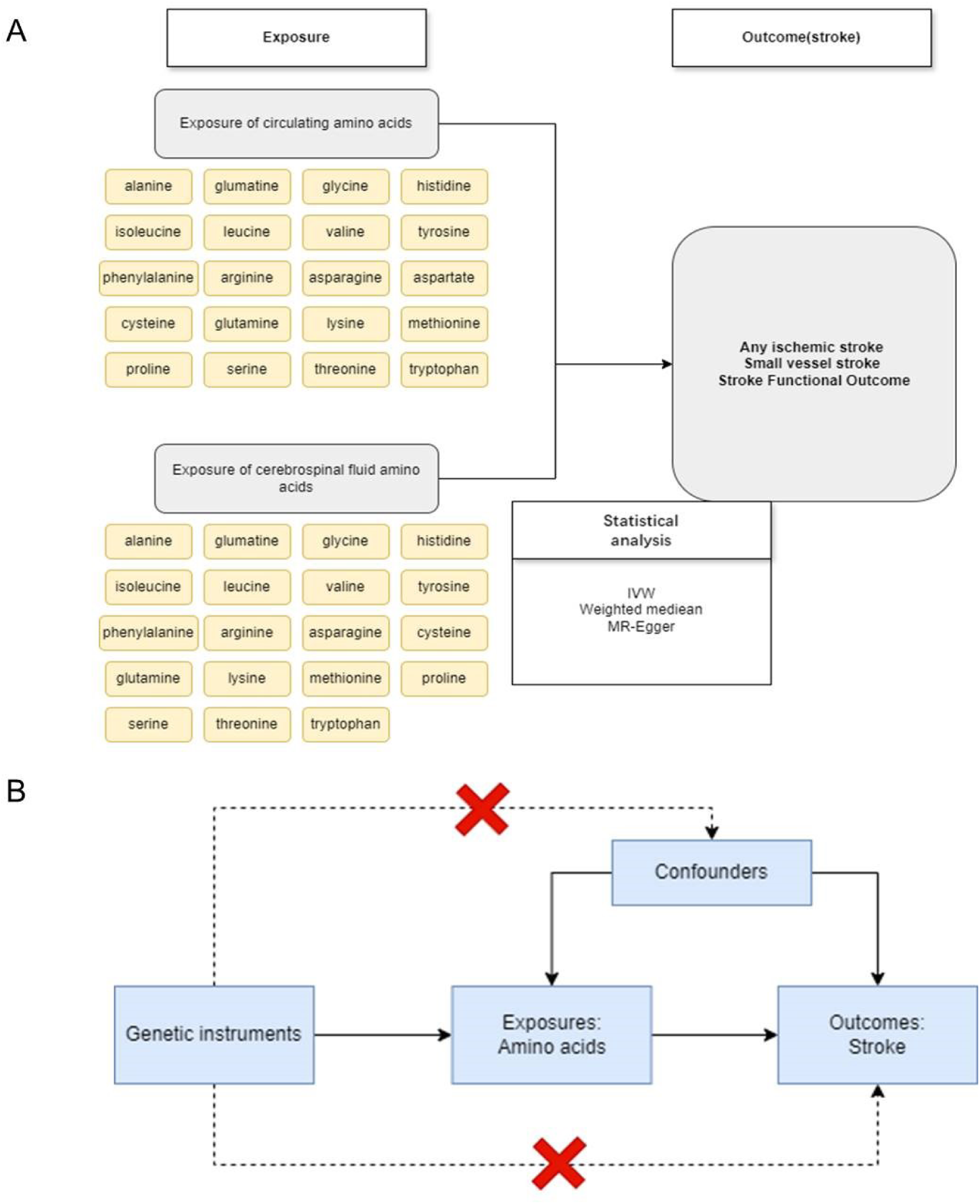
An overview of the mendelian randomization study design for the association between amino acids and the risk of ischemic stroke, small vessel stroke, and ischemic stroke. The assumption is that 1) instrumental variables are associated with amino acid levels, 2) instrumental variables are not associated with confounders, 3) and instrumental variables affect ischemic stroke or its subtypes only through the effects on amino acid levels. Abbreviation: SNP, single-nucleotide polymorphism; IVW, inverse-variance weighted.

### Data sources on the amino acids

We selected 20 common circulating amino acids, including alanine, arginine, asparagine, aspartate, cysteine, glutamate, glutamine, glycine, histidine, isoleucine, leucine, lysine, methionine, phenylalanine, proline, serine, threonine, tryptophan, tyrosine and valine. Summary statistics for glutamine, glycine, histidine, leucine, valine, phenylalanine levels, and tyrosine levels were from the study conducted in 2023(Davyson et al. 2023). In this study, a GWAS for amino acids (AAs) was conducted using data from approximately 88000 individuals. Summary statistics for the other 12 AAs were from the study conducted in European populations in 2023(Chen et al. 2023), which contained approximately 8,245 participants. The CSF amino acid data utilized in our study was derived from a 2021 study involving 291 individuals of European ancestry (Panyard et al. 2021). Overall, a total of 20 amino acids including circulating amino acids and 19 cerebrospinal fluid amino acids as exposure (Figure 1.A).

### Instrumental variables selection

In this study, we initially applied a stringent threshold of p < 5 × 10⁻⁸, pairwise linkage disequilibrium r^2^<0.001 within a 10,000 kb window, to screen for single nucleotide polymorphisms (SNPs) strongly associated with each circulating amino acid, stroke, and functional outcomes following ischemic stroke (IS). However, when certain circulating amino acids were considered as exposures, only a few SNPs met this criterion. Consequently, we broadened our approach to include SNPs that reached significance at a locus-wide threshold (p < 5 × 10^⁻⁶^, r^2^<0.001, kb = 10,000). These SNPs were selected as instrumental variables (IVs) to enhance both the depth and sensitivity of the subsequent MR analyses.

The potency of each genetic instrument was quantified by the F-statistic, calculated through the formula 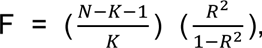 where R^2 represents the proportion of phenotypic variance explained by the IV, and N is the effective sample size obtained from the GWAS. Only SNPs with an F-statistic exceeding ten were retained for inclusion in the MR analysis to ensure strong and reliable estimates of genetic association (Bottigliengo et al. 2022), thus precluding the risk of weak instrument bias for F-statistics greater than 10(Burgess and Thompson 2011).

As a final measure to enhance the validity of our findings, SNPs with palindromic sequences that could introduce allele determination ambiguity were systematically excluded. This step was critical to guarantee the precision and trustworthiness of our results.

### Data sources on the stroke and its functional outcome

Stroke is classified based on the clinical criteria defined by the World Health Organization (WHO) and the tenth edition of the International Classification of Diseases (ICD-10) (Thayabaranathan et al. 2022). The MEGASTROKE consortium carried out an extensive investigation within a cohort consisting of 446,696 individuals of European ancestry. This cohort included 40,585 cases classified as any stroke (AS) and was contrasted with a control group comprising 406,111 individuals. Within the specific category of any ischemic stroke (AIS), the consortium identified a total of 34,217 AIS cases and 5,386 cases of small vessel stroke (SVS).

In addition to the aforementioned data, we also scrutinized GWAS information pertinent to functional outcomes following an ischemic stroke as reported by the GISCOME network, which included 6,021 patients of European descent (Söderholm *et al*. 2019). Functional outcomes three months post-stroke were assessed using the modified Rankin Scale (mRS), with a score range of 0–2 signifying better outcomes (in 3741 patients), and 3–6 indicating poorer outcomes (in 2280 patients). The primary MR analysis utilized results that had been corrected for variables such as age, sex, ancestry, and initial stroke severity. Additionally, for the sake of comparison, supplementary MR analyses were conducted using GWAS data not adjusted for stroke severity.

### Statistical analysis

This investigation applied inverse variance weighting (IVW) as the primary statistical approach for MR analysis. In this analysis, associations were initially identified through a nominal p-value threshold of less than 0.05. The heterogeneity among genetic instruments was evaluated using Cochran’s Q statistic(Greco et al. 2015). A random-effects IVW model was employed when heterogeneity was present; otherwise, a fixed-effects IVW model was applied.

The FDR approach was used to correct for multiple testing, applied to the p-values from the IVW random-effect model (Korthauer et al. 2019). An association was considered significant if the p value was less than 0.05 and deemed suggestive if the unadjusted p-value was less than 0.05.

### Sensitivity and reverse MR analyses

In the sensitivity analyses, we employed various MR methods to assess the robustness of our findings. These methods included penalized IVW (Rees et al. 2019), maximum likelihood (Hemani et al. 2018), MR-Robust Adjusted Profile Score [MR-RAPS] (Zhao et al. 2019) and MR-Egger regression (Bowden et al. 2015). The penalized IVW method adjusts the weights of SNPs with pleiotropy (Rees *et al*. 2019). The maximum likelihood method provides reliable estimates despite measurement errors in the SNP-exposure effect (Hemani *et al*. 2018). MR-RAPS addresses biases from horizontal pleiotropy and weak instruments (Zhao *et al*. 2019), while MR-Egger regression evaluates the average pleiotropic effects across all SNPs via the intercept term (Bowden *et al*. 2015).

Using the same selection criteria mentioned above, we selected SNPs from GWAS on ischemic stroke, small vessel stroke, and ischemic stroke functional outcomes for bidirectional Mendelian randomization analysis to explore potential reverse causal relationships. We obtained comprehensive summary statistics data from previous studies and conducted analyses primarily using the MR-IVW method. The results showed statistical significance with a P-value less than 0.05.

Results were presented as odds ratios (ORs) with 95% confidence intervals (CIs). Amino acids-stroke association was considered robust if it met multiple criteria: FDR-corrected IVW significance, consistency across MR methods, F-statistics > 10 for all IVs. Analytical procedures were conducted using R software, version 4.4.0, with the employment of “TwosampleMR” and “MendelianRandomization” packages for comprehensive analysis.

### Power calculation

To assess the statistical power for detecting causal relationships in MR analyses, we employed the mRnd online calculator (https://shiny.cnsgenomics.com/mRnd/)(Brion et al. 2013). This resource utilizes asymptotic theory to project the power of MR studies to identify causal effects using instrumental variables (IVs). Our power calculations were conducted with a Type I error rate set at 0.05, incorporating critical parameters including the R² attributable to the IVs, the case proportion from the cancer GWAS, and the odds ratio (OR) from MR analyses executed via the Inverse Variance Weighted (IVW) method.

## Results

### SNP Screening and Instrumental Variable Analysis

In order to ensure results of SNPs for subsequent MR analysis, a threshold of p < 5 × 10 -6 was selected when screening SNPs related to each amino acids and stroke, along with functional outcome following IS. The F-statistics for SNPs of the amnio acids and CSF amino acid were 19.52–14621.42. The F-statistic for all used instrumental variables was greater than 10, indicating no weak instrumental variable bias.

### Effects of amino acids on stroke and functional outcomes following IS

In this study, a total of 1258 SNPs were used as genetic instruments for amino acids, with details provided in Table S2. The F-statistics for these instrumental variables were above 10, indicating the absence of weak instrument bias (Table S2). The main analysis, conducted using IVW models, examined the association of amino acids with stroke and its functional outcomes.

As illustrated in Figure 2, the primary IVW MR analysis demonstrated that genetically determined high isoleucine levels were associated with increased risks of ischemic stroke (OR per standard deviation [SD] increase: 1.07; 95% CI: 1.01-1.15; P=0.03). Similarly, high glutamine levels were associated with increased risks of SVS (OR per SD increase: 1.20; 95% CI: 1.01-1.42; P=0.04), and high alanine levels were linked to increased risks of functional outcome after IS (OR per SD increase: 1.44; 95% CI: 1.06-1.97; P=0.02).

**Figure 2.**
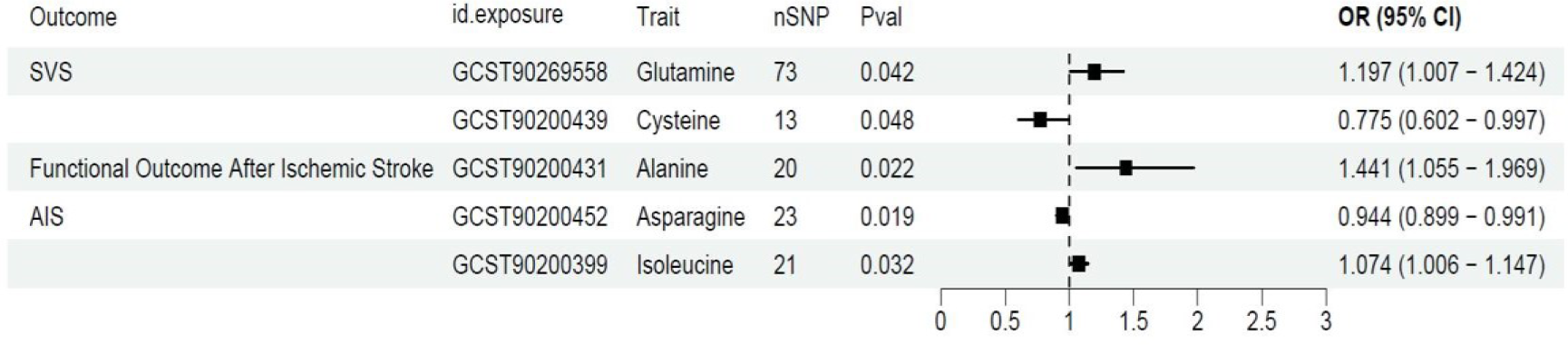
The associations between genetically determined amino acids and the risks of ischemic stroke, small vessel stroke, and ischemic stroke functional outcomes were analyzed using the inverse-variance weighted MR analysis. Abbreviations: OR, odds ratio; 95% CI, 95% confidence interval.

Conversely, high asparagine levels were associated with decreased risks of AIS (OR per SD decrease: 0.94; 95% CI: 0.90-0.99; P=0.02), and high cysteine levels were linked to decreased risks of SVS (OR per SD decrease: 0.78; 95% CI: 0.60-1.00; P=0.05). The associations between each instrumental variant for amino acid levels and the risks of ischemic stroke, small vessel stroke, and their functional outcomes are presented in Figure 4.

**Figure 3.**
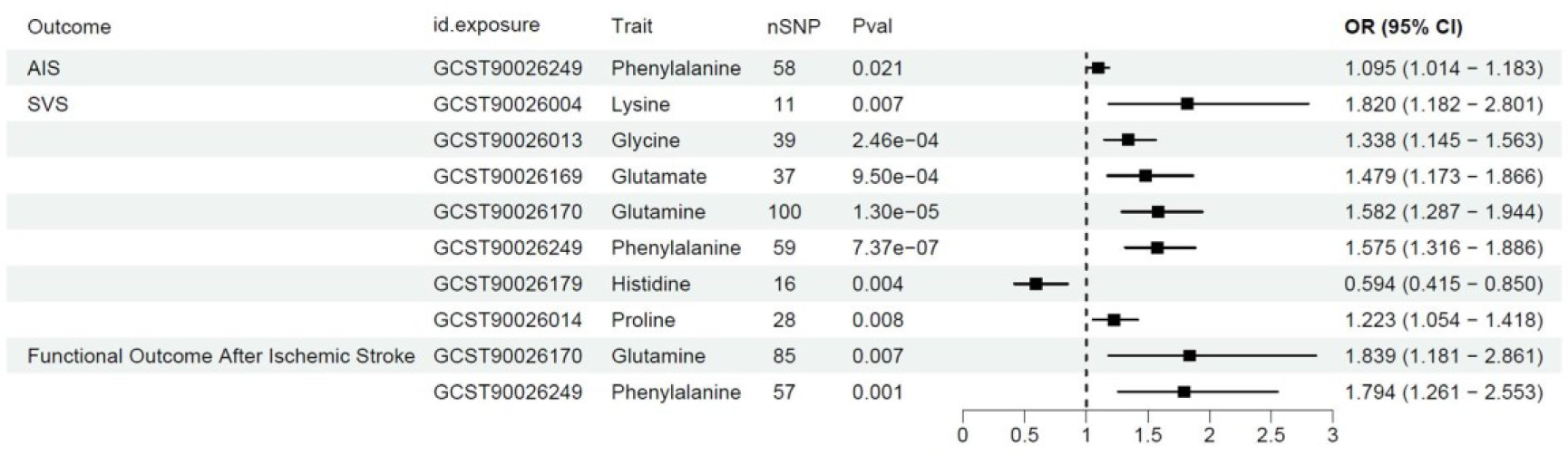
The associations between genetically determined cerebrospinal fluid (CSF) amino acids and the risks of ischemic stroke, small vessel stroke, and ischemic stroke functional outcomes were analyzed using the inverse-variance weighted MR analysis. Abbreviations: OR, odds ratio; 95% CI, 95% confidence interval.

**Figure 4.**
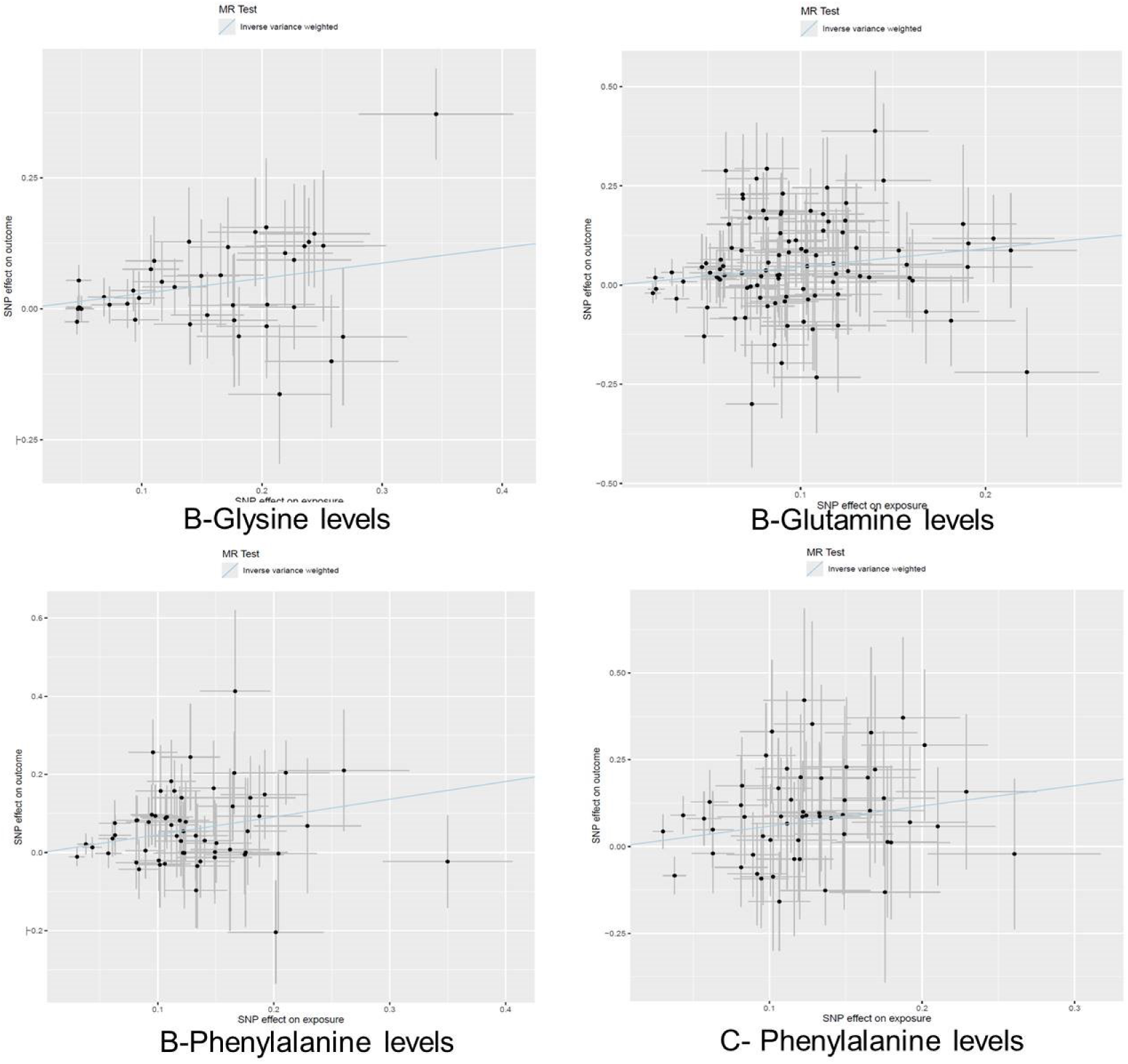
Associations between genetic instruments of cerebrospinal fluid (CSF) amino acid and the risk of ischemic stroke, small vessel stroke and ischemic stroke functional outcome. The line indicates the estimate for the associations of with ischemic stroke, small vessel stroke and stroke functional outcome using inverse-variance weighted method. Circles indicate associations of each genetic variant related to amino acid levels with the risk of ischemic stroke and cardioembolic stroke. Error bars genetic indicate 95% confidence interval. A, Any ischemic stroke; B, Small vessel stroke; C, Functional outcome after ischemic stroke.

However, after FDR correction, no association was found between amino acids and stroke or stroke functional outcome, indicating that any positive results may have been reduced to only suggestive of a positive association.

### Effects of CSF amino acids on stroke and functional outcomes following IS

As illustrated in Figure 3, the primary IVW MR analysis demonstrated that genetically determined high phenylalanine levels were associated with increased risks of ischemic stroke (OR per standard deviation [SD] increase: 1.10; 95% CI: 1.01-1.18; P=2.06E-02) and small vessel stroke (SVS) (OR per SD increase: 1.58; 95% CI: 1.32-1.89; P=7.37e-07). Similarly, high levels of lysine (OR per SD increase: 1.82; 95% CI: 1.18-2.80; P=6.52E-03), glycine (OR per SD increase: 1.34; 95% CI: 1.14-1.56; P=2.46E-04), proline (OR per SD increase: 1.22; 95% CI: 1.05-1.42; P=7.88E-03), glutamate (OR per SD increase: 1.48; 95% CI: 1.17-1.87; P=9.50E-04), and glutamine (OR per SD increase: 1.58; 95% CI: 1.29-1.94; P=1.30E-05) were associated with increased risks of SVS. Additionally, high glutamine (OR per SD increase: 1.84; 95% CI: 1.18-2.86; P=6.96E-03) and phenylalanine (OR per SD increase: 1.79; 95% CI: 1.26-2.55; P=1.15E-03) levels were linked to increased risks of worse stroke functional outcomes.

Conversely, high histidine levels were associated with decreased risks of SVS (OR per SD decrease: 0.59; 95% CI: 0.41-0.85; P=4.44E-03). The associations between each instrumental variant for amino acid levels and the risks of ischemic stroke, small vessel stroke, and their functional outcomes are presented in Figure 2.

After FDR correction, we observed 4 CSF amino acids (glycine, glutamate, glutamine, and phenylalanine) with significant causative correlations to SVS and CSF phenylalanine levels with functional outcomes following stroke.

### MR sensitivity analyses results

We conducted sensitivity analyses of amino acids using a series of Mendelian Randomization (MR) methods to assess the robustness of our findings (Table 1). Genetically determined glycine was positively associated with the risk of small vessel stroke (SVS) in the sensitivity analyses using the penalized inverse variance weighted (IVW) method (OR per standard deviation (SD) increase: 1.26; 95% CI: 1.07-1.48; P = 6.00E-03), MR-RAPS method (OR per SD increase: 1.36; 95% CI: 1.15-1.59; P = 2.13E-04), and maximum likelihood method (OR per SD increase: 1.36; 95% CI: 1.16-1.59; P = 1.65E-04).

**Table 1.**
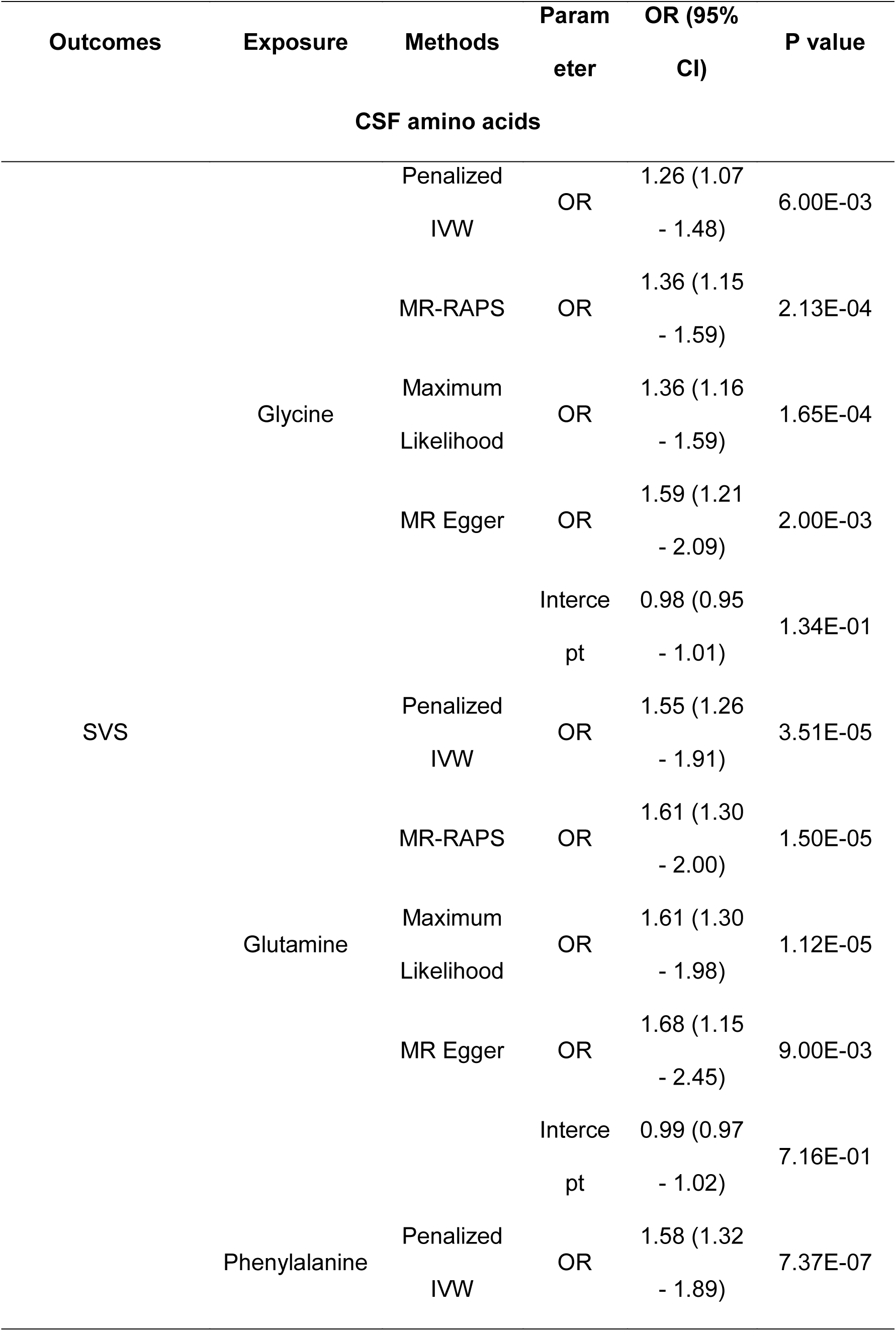
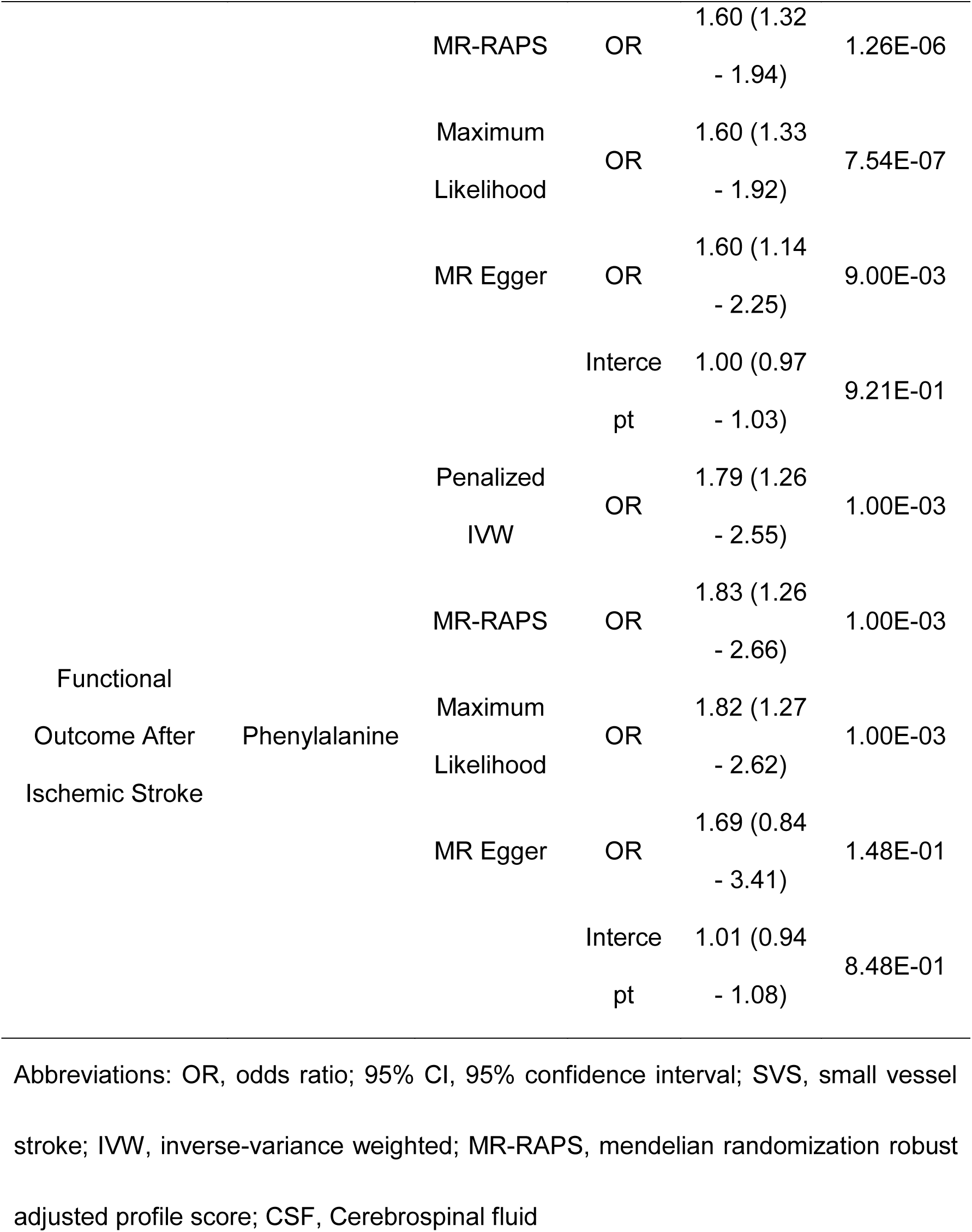
Sensitivity analyses for the associations of genetically determined amino acids with the risk of stroke and stroke functional outcome.

Genetically determined glutamine was also positively associated with the risk of SVS in the sensitivity analyses using the penalized IVW method (OR per SD increase: 1.55; 95% CI: 1.26-1.91; P = 3.51E-05), MR-RAPS method (OR per SD increase: 1.61; 95% CI: 1.30-2.00; P = 1.50E-05), and maximum likelihood method (OR per SD increase: 1.61; 95% CI: 1.30-1.98; P = 1.12E-05).

Furthermore, genetically determined phenylalanine was positively associated with the risk of SVS in the sensitivity analyses using the penalized IVW method (OR per SD increase: 1.58; 95% CI: 1.32-1.89; P = 7.37E-07), MR-RAPS method (OR per SD increase: 1.60; 95% CI: 1.32-1.94; P = 1.26E-06), and maximum likelihood method (OR per SD increase: 1.60; 95% CI: 1.33-1.92; P = 7.54E-07).

Additionally, genetically determined phenylalanine was positively associated with the risk of poor functional outcome after ischemic stroke in the sensitivity analyses using the penalized IVW method (OR per SD increase: 1.79; 95% CI: 1.26-2.55; P = 1.00E-03), MR-RAPS method (OR per SD increase: 1.83; 95% CI: 1.26-2.66; P = E-03), and maximum likelihood method (OR per SD increase: 1.82; 95% CI: 1.27-2.62; P = 1.00E-03).

### Reverse MR shows no reverse causation

In reverse MR analyses, none of these associations showed the possibility of reverse causation (all P > 0.05) (detailed in Table S4).

## Discussion

To our knowledge, this is the first study to use bidirectional two-sample Mendelian randomization (MR) to investigate the causal relationship between plasma and cerebrospinal fluid (CSF) amino acid levels and ischemic stroke, its subtypes, and patient prognosis. Based on genome-wide association studies (GWAS), we found that several CSF amino acids are associated with the risk of SVS or mRS. For example, high CSF levels of glutamate, glutamine, glycine, and phenylalanine are associated with an increased risk of SVS, and high phenylalanine levels are also positively correlated with poor stroke outcomes. These findings suggest that CSF amino acid levels play a key role in the development of SVS and patient prognosis.

In recent years, several observational studies have shown that amino acid levels in plasma and CSF are significantly associated with the risk of ischemic stroke and its subtypes. In a large multicenter cohort study involving 3,486 ischemic stroke patients, elevated plasma levels of glutamate, aspartate, and γ-aminobutyric acid, and reduced levels of glycine were associated with poor outcomes after ischemic stroke(Zhu et al. 2023). Another cohort study involving seven cohorts and 38,797 participants found that circulating amino acids such as histidine were associated with a reduced risk of stroke, while high levels of pyruvate and phenylalanine were significantly associated with an increased risk of ischemic stroke (Vojinovic et al. 2021).

However, traditional observational studies have some limitations, such as difficulty controlling for all potential confounding factors and the inability to clearly establish the temporal sequence of exposure and outcome, making it difficult to determine the direction of causality. Therefore, to avoid these biases and reasonably infer the potential causal relationship between amino acid levels and ischemic stroke, our MR study used genetic variations as instrumental variables to assess the causal relationship between plasma and CSF amino acids and stroke risk and prognosis.

This study focused on the analysis of amino acid levels in the cerebrospinal fluid (CSF), which directly reflects the metabolic state of the central nervous system. We found that high CSF phenylalanine levels are significantly associated with the risk of SVS and poor stroke prognosis. Elevated phenylalanine levels may lead to disease onset and adverse outcomes by affecting neurotransmitter metabolism and neuronal function. In the MCAO rat model, phenylalanine levels were significantly elevated, and inhibiting phenylalanine could reduce reperfusion injury (Chun-bing 2018). A clinical study also found that phenylalanine levels were elevated in the serum of IS patients, and phenylalanine levels could serve as a biomarker for diagnosing IS (Ormstad et al. 2016). High concentrations of phenylalanine can affect Ca2+ and ERK levels and inhibit neuronal synapse growth (Hartwig et al. 2006).

Another finding of our study is that high levels of glutamate, glutamine, and glycine in CSF increase the risk of SVS. Glutamate and its precursor glutamine, as abundant amino acids in the body, participate in various biological processes. Elevated levels of these two amino acids can induce excitotoxicity leading to neuronal death (Matsumoto et al. 1993; Wang et al. 2021), thereby exacerbating stroke injury (He et al. 2024). Glycine, as a co-agonist of NMDA receptors, can enhance glutamate-dependent calcium influx into cells (Rothman and Olney 1986; Siesjö and Bengtsson 1989; Takagi et al. 1993), aggravating neuronal death.

Clinical studies have shown that CSF amino acids are closely related to stroke occurrence or progression. Multiple studies on AIS patients have found significantly elevated concentrations of glutamate, glutamine, and glycine in both plasma and CSF (Castillo et al. 1996; Castillo et al. 1997; He *et al*. 2024). In focal cerebral ischemia animal models, glycine expression levels were also significantly elevated. The differences in amino acid levels between CSF and serum are more pronounced, reflecting that under ischemic and hypoxic conditions, the blood-brain barrier may be disrupted, leading to increased permeability.

Our study results have important public health and clinical implications. Firstly, this study is the first to explore the genetic determination of circulating and CSF amino acid levels in relation to stroke and its subtypes and outcomes using MR analysis, which may provide new clues for stroke prevention. Amino acid levels can serve as biomarkers to identify high-risk individuals, assess patient prognosis, enable proactive monitoring, and early intervention. Adjusting amino acid intake through diet or medication offers new strategies for stroke prevention and treatment.

This MR study has several advantages. Firstly, bidirectional MR analysis avoids reverse causality and confounding bias. Secondly, we used a large sample GWAS database for comprehensive evaluation, and validated the robustness and reliability of the results through various methods and sensitivity analyses.

However, this study has some limitations. Firstly, the pleiotropy of instrumental variables may bias the results. Secondly, the study population is mainly of European descent, limiting the generalizability of the results to other ethnic groups. Finally, although our study provides potential genetic determination of amino acid levels in relation to ischemic stroke and its subtypes, these findings still require further clinical validation. Future research should be conducted in larger and more diverse populations, combined with actual clinical data, to confirm our findings and explore the feasibility and effectiveness of their clinical application.

In conclusion, this study emphasizes the potential role of amino acids as targets for stroke intervention. While further research is needed to translate these findings into clinical practice, it lays the groundwork for understanding the role of amino acids in stroke and developing innovative treatment strategies. These insights support the importance of metabolic health in stroke prevention and recovery, providing new avenues for research and therapeutic development.

## Conclusions

This study used bidirectional Mendelian randomization to show that elevated CSF amino acids like phenylalanine, glutamate, glutamine, and glycine are linked to higher risks of small vessel stroke (SVS) and poor outcomes. These amino acids could serve as biomarkers for stroke risk and prognosis, offering new prevention and treatment strategies. Further validation in diverse populations is needed.

## Data Availability

We thank the authors and participants of all GWASs that we have used for making their results publicly available, which provided summary statistics data for making these analyses. Full acknowledgement and funding statements for each of these resources are available via the relevant cited reports.

## Acknowledgements

We thank the authors and participants of all GWASs that we have used for making their results publicly available, which provided summary statistics data for making these analyses. Full acknowledgement and funding statements for each of these resources are available via the relevant cited reports. This study was supported by the Basic Frontier Innovation Cross Program of Suzhou Medical College (YXY2304057).

## Competing interests

The authors declare that there is no conflict of interests.

## Notes

### Competing Interest Statement

The authors have declared no competing interest.

